# Decontamination of filtering facepiece respirators in primary care using medical autoclave

**DOI:** 10.1101/2020.04.27.20066654

**Authors:** Ralf E. Harskamp, Bart van Straten, Jonathan Bouman, Bernadette van Maltha - van Santvoort, John J. van den Dobbelsteen, Joost R.M. van der Sijp, Tim Horeman

## Abstract

**Objective:** There are widespread shortages of personal protective equipment as a result of the coronavirus disease 2019 (COVID-19) pandemic. Reprocessing filtering facepiece respirators may provide an alternative solution in keeping health care professionals safe.

**Design:** prospective, bench-to-bedside

**Setting:** A primary care-based study using filtering facepiece particles (FFP) type 2 respirators without exhalation valve (3M Aura 1862+, Maco Pharma ZZM002), FFP2 respirators with valve (3M Aura 9322+ and San Huei 2920V), and valved FFP type 3 respirators (Safe Worker 1016).

**Interventions:** All masks were reprocessed using a medical autoclave (34-minute total cycle time of steam sterilization, with 17 minutes at 121°C) and subsequently tested up to 3 times whether these decontaminated respirators retained their integrity (seal check, pressure drop) and ability to filter small particles (0.3-5.0μm) in the laboratory using a particle penetration test.

**Results:** We tested 32 respirators, and 63 samples for filter capacity. All 27 FFP-2 respirators retained their shape, whereas half of the sterilized FFP-3 respirators (Safe Worker 1116) showed deformities and failed the seal check. The filtering capacity of the 3M Aura 1862 was best retained after 1, 2, and 3 sterilization cycles (0.3μm: 99.3±0.3% (new) versus 97.0±1.3, 94.2±1.3% or 94.4±1.6, p<0.001). Of the other FFP-2 respirators, the San Huei 2920V had 95.5±0.7% at baseline versus 92.3±1.7% versus 90.0±0.7 after one- and two-time sterilization, respectively (p<0.001). The tested FFP-3 respirator (Safe Worker 1016) had a filter capacity of 96.5±0.7% at baseline and 60.3±5.7% after one-time sterilization (p<0.001). Breathing and pressure resistance tests indicated no relevant pressure changes between respirators that were used once, twice or thrice.

**Conclusion:** This study shows that selected FFP2-type respirators may be reprocessed for use in primary care, as the tested masks retain their shape, ability to retain particles and breathing comfort after decontamination using a medical autoclave.

**Strengths and limitations of this study:**

- Pragmatic use of autoclave to sterilize and reuse filter facepiece respirators
- Combining clinical and laboratory findings to evaluate the safety in terms of shape, ability to retain particles and breathing comfort
- The study is limited in sample size and restricted to selected FFP-2 and FFP-3 respirators
- The study is a first of its kind in primary care settings and thus unvalidated
- The study does not provide "hard" clinical evidence in terms of a randomized trial (i.e. reprocessed mask versus usual care)

## Introduction

General practitioners (GP) are often the first to evaluate patients with (suspected) coronavirus disease 2019 (COVID-19). This is particularly true in countries where GPs have a gatekeeping role. Given the risk of person-to-person spread this necessitates the need to wear personal protective equipment. [1,2] Unfortunately, most health care facilities are running dangerously low on this equipment. [1,2] In the United States, these critical shortages have resulted in downgrading from respirators to surgical masks and now even resort to home-made cloth-masks. [1] Access to adequate supplies is crucial to preventing transmission of pathogens, especially in resource-limited settings. [3] Reports across several countries found that healthcare workers are more at risk of catching SARS-CoV2 as well as at higher risk of severe COVID-19, possibly due to exposure to higher viral load. [4] The outbreak of COVID-19 in Italy showed that inadequate access to protective equipment is one of the reasons why healthcare workers, and particularly GPs, experienced high rates of infection. [2] Aside from the direct health effects, absenteeism from illness may also negatively affect the health system’s capacity to adequately respond to the COVID-19 pandemic. Moreover, it makes healthcare workers feel unsafe and unprotected, which undermines morale as shown in a report on England’s National Health Service health workers experiences. [5]

One of the possible (short-term pragmatic) solutions could be the reuse of equipment, and in particular that of respirators. To reuse a mask or respirator, it should be sterilized first. The method applied should: 1) kill the SARS-CoV-2 virus (diminish the viral load); 2) keep the mask’s protective properties (largely) intact, in terms of filter and fit. In primary care, the medical autoclave is normally used to sterilize surgical instruments. The process of pressurized moist heat destroys microorganisms by the irreversible coagulation and denaturation of enzymes and structural proteins, and has been shown to be effective in respirators contaminated with other viruses, such as H1N1 influenza virus. [2, 6, 7] However, the question is whether the respirator’s protective properties in terms of filter function and fit will remain intact when exposing the respirator to steam. We therefore set out to study whether the process of steam sterilization negatively affects the protective properties of commonly used respirators which are designed to protect the wearer against the inhalation of both droplets and particles suspended in the air.

## Methods

We reported our findings according to the Better reporting of interventions: template for intervention description and replication (TIDieR) checklist and guide, as well as the general principles of reporting a study using the directions provided by the journal. [8]

### Study design and setting

The study involved the evaluation of available filtering respirators used to evaluate suspected COVID-19 patients in the Holendrecht Medical Center, in Amsterdam, the Netherlands. For high-risk patients, the center provides GPs with filtering facepiece particles (FFP) type 2 or type 3 for personal protection. For the current study, worn respirators were used for reprocessing using a medical autoclave. After the autoclave procedure the respirators were visually inspected for deformity by two clinical investigators, followed by a seal check. The masks were subsequently marked and sent by courier to the GreenCycl testing laboratory in Utrecht, the Netherlands. At this facility, the sterilized respirators were tested by two laboratory scientists for their filter capacity. For comparison, the results ware compared with the filter capacity of unused, brand-new respirators that were used as a reference. Moreover, a pressure drop test was performed to evaluate whether the breathing resistance altered by the process of sterilization.

### Sterilization process

The masks were sterilized for multiple cycles using a cylindrical chamber tabletop autoclave (Kronos S18, release: E.5.47a, Newmed, Quattro Castella, Italy). This type of vacuum autoclave is typically designed for general practitioner and dental practices and has pre-programmed cycles. The size as well as the cycle times differ from autoclaves typically used in hospitals, which are larger and have longer cycle times, however with comparable peak times regarding sterilization. The Kronos S18 autoclave holds a capacity of 18L or 4 respirators. The autoclave has specific programs for “solid made of rubber and delicate solids”, which includes respirators. The sterilization program we used involved a 34-minute cycle, of which the first 12 minutes of the cycle involved preheating, followed by 17 minutes steam sterilization at a temperature of 121 degrees of Celsius, and finished with a 5-minute drying process.

### Visual inspection, breathing resistance, and user seal check

After sterilization the respirators were checked for visual deformities of the mask as well as the elastic straps. Subsequently, the respirators were put on to evaluate whether breathing felt normal, followed by the performance of a user seal check. A negative pressure user seal check was used for all respirators in which the clinical investigator inhaled sharply while blocking the paths for air to enter the facepiece. A successful check is when the facepiece collapsed slightly under the negative pressure that was created with this maneuver. For respirators without an exhalation valve the investigator also performed a positive pressure check by exhaling gently while blocking the paths for air to exit the facepiece. A successful check is when the facepiece was slightly pressurized before increased pressure causes outward leakage. [9]

### Particle penetration test

At the testing laboratory two independent researchers from the Delft University of Technology tested the masks using a dry particle penetration test setup (*Figure 1A*)[10] The equipment involved a SOLAIR 3100 particle counter (Lighthouse Worldwide Solutions Inc, Fremont, CA). The particles are counted within the machine via a tube that is connected to a particle chamber to which the respirator is secured. The transparent lid presses the mask such that it prevents material buckling and creates an airtight seal that only allows air to pass through the material. Before each measurement, a benchmark test is conducted with 28 Liters of surrounding air that is sucked through the particle chamber into the particle counter. During this measurement no mask is installed. During the test measurement, a mask is installed on the particle chamber. Therefore, the 28 liter of surrounding air is sucked through the filter material of the mask and the remaining particles are counted in the categories of 0.3, 0.5, 1.0 and 5.0 microns. The measurements are compared and the filtering capacity derived based on the difference in the readings compared to the benchmark test. A lower number of particles counted after filtering in relation to the benchmark test would indicate better filtering performance. [11] The system setup is more conservative than the NEN-149 standard, which means the resulting filter capacity percentages cannot be translated directly to the known FFP1, 2 and 3 standards. However, as the filter capacity of a new mask is known, the measurement results do show the remaining filter capacity and therefore indicates whether a mask type deteriorates after steam sterilization.

**Figure 1.**
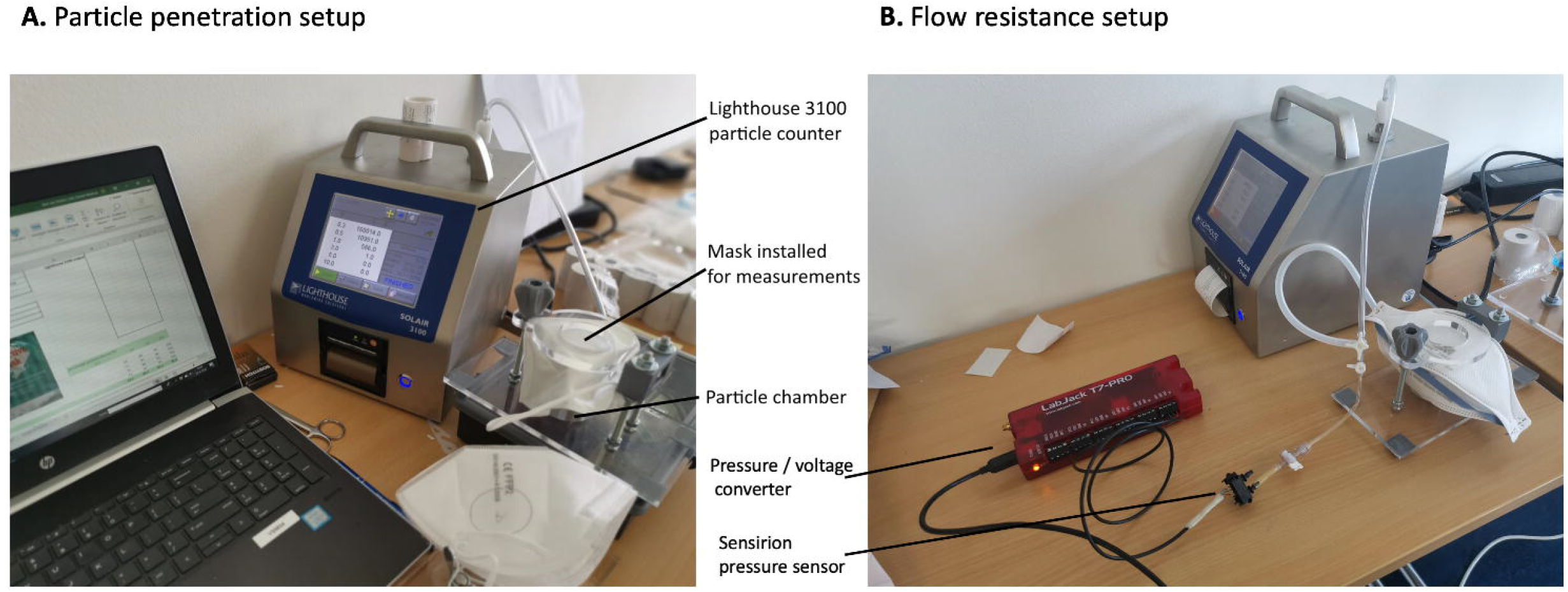
Panel A (left side) illustrates the measurement setup used to measure the particle penetration capacity of different respirators. The particle chamber is connected to the Lighthouse 3100 with a custom connecter and 5 mm tube. An adjustable removable transparent lid is used to press the filter material of a mask airtight on the rim of the open particle chamber for accurate measurements. Panel B (right side) shows an expansion of the setup with an additional pressure sensor and pressure-to-voltage converter.

### Flow resistance

The setup was expanded with an additional pressure sensor (SDP 816-500Pa Sensirion #1230319) and flow adjustment valve in order to investigate whether the pressure delta over the mask material changed after sterilization cycles. (*Figure 1B*) The Sensirion pressure sensor was connected with a T-piece between the particle chamber and Lighthouse 3100. An additional valve was used to adjust the input pressure within range of the sensor. A LabJack T7 analogue input device was used to convert the output from the pressure sensor to an output voltage of 0 to 5 Volt. An output value of 5V was representing 500Pa and set as 100% of input pressure. the atmospheric input pressure of 2.42 Volt was defined as 0% output. Measurements were conducted with a constant air speed of 20.7 meter per seconds at the opening of the particle chamber.

### Outcomes of interest

The outcomes of interest involved: 1) signs of deformity of the respirator, which was performed by visual inspection; and 2) the percentage of filtered particles with a diameter of 0.3 μm. This diameter is clinically relevant, given that to meet the FFP-2 standards a mask should filter 94% of all 0.3 μm particles, whereas 99% of these particles should be filtered to meet the FFP-3 standard.

### Statistical analysis

The study involved descriptive analyses, with numbers and percentages, and comparisons were performed using an alpha of 0.05 for statistical significance. The findings of the filter tests are visually displayed in Box plots, and presented as mean and standard deviation. We used JASP statistical software (version 0.10.2, University of Amsterdam, the Netherlands).

### Patient and Public Involvement

Patients and/or the public were not involved in the design, or conduct, or reporting, or dissemination plans of our research.

## Results

We obtained 32 respirators, of which 27 were used during consultation or high-risk home visits of COVID-19 suspected patients at the Holendrecht Medical Center in March/April 2020. The facemasks were FFP-2 respirators (3M Aura 1862+, Maco Pharma ZZM002), FFP2 respirators with exhalation valve (3M Aura 9322+ and San Huei 2920V), or FFP3 respirators (Safe Worker 1016). The 27 used respirators (including 4 FFP3 respirators) underwent sterilization, with the remaining 5 serving as a reference (as they did not to undergo sterilization).

### Visual inspection, breathing resistance, and user seal check

After the sterilization process all FFP-2 respirators retained their shape and were without visible damage. When fitting, the elastic bands of all masks still functioned normally, with no difference from non-sterilized masks in terms of breathing resistance. The seal checks also did not reveal significant air leakage suggesting poor fit. However, unlike the FFP-2 respirators, two out of the four FFP3 respirators (50%) showed signs of deformation, with a crumbled appearance, and failed seal check test.

### Filter capacity of sterilized respirators

For the particle penetration test, a total of 63 samples were tested from 32 respirators. The results of the filter capacity for 0.3 microns are illustrated in *Figure 2* and for larger particles are displayed in *Table 1*. Of the tested FFP-2 respirators we found that the 3M Aura 1862+ remained close to its original filtering capacity after one-, two-time, and three-time sterilization (0.3μm: 99.3±0.3% versus 97.0±1.3, 94.2±1.3% or 94.4±1.6, respectively, p<0.001). The 3M Aura 9322+ (with valve) had a filter capacity of 96.8±0.2% without sterilization versus 91.0±1.4% and 77.5±2.1% after one- or two-time sterilization (p<0.001). The Maco Pharma ZZM002 FFP-2 mask did not have a reference mask, but after one- and two-time sterilization the filter capacities were 89.3±3.9% and 86.6±2.6%, respectively. The San Huei 2920V respirator had 95.5±0.7% at baseline versus 92.3±1.7% versus 90.0±0.7 after one- and two-time sterilization (p<0.001). Finally, the tested FFP-3 respirator (Safe Worker 1016) had a filter capacity of 96.5±0.7% at baseline and 60.3±5.7% after one-time sterilization (p<0.001).

**Figure 2.**
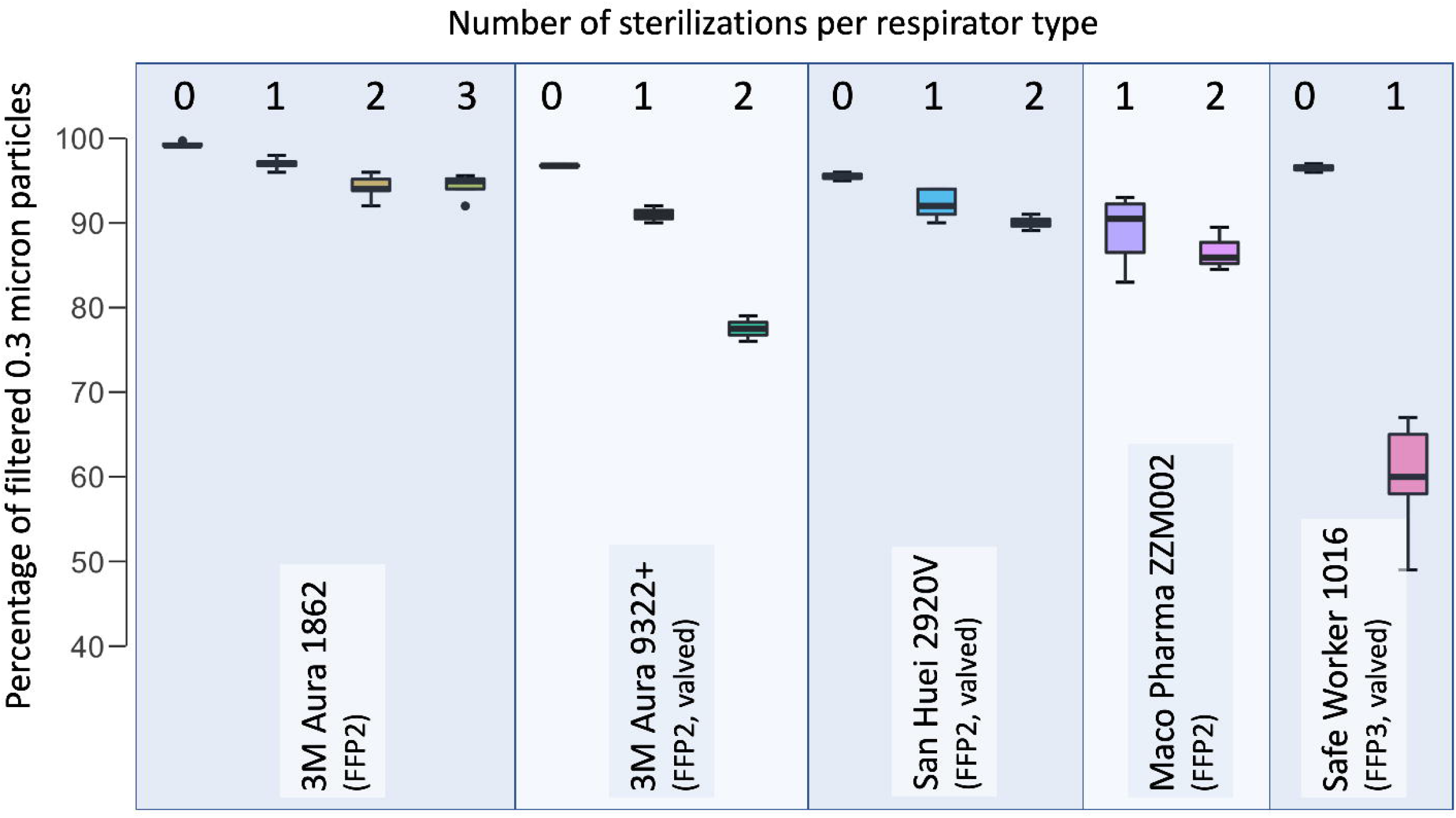
Filter quality of autoclave-decontaminated respirators (retainment of 0.3μm particles) of unused, and one-time, two-time, and three-time autoclave sterilization

**Table 1.** Filter capacity of the tested respirators by particle size

| Respirator | Condition (new/sterilized) | No of samples | 0.3 $\mu\text{m}$ | 0.5 $\mu\text{m}$ | 1.0 $\mu\text{m}$ | 5.0 $\mu\text{m}$ |
| --- | --- | --- | --- | --- | --- | --- |
| 3M Aura 1862+ | New | 4 | 99.3±0.3 | 99.7±0.0 | 99.8±0.1 | 99.9±0.2 |
| | 1x | 4 | 97.0±0.8 | 99.0±0.0 | 99.0±0.5 | 100±0.0 |
| | 2x | 8 | 94.2±1.3 | 97.4±0.5 | 98.9±0.3 | 99.9±0.1 |
| | 3x | 4 | 94.4±1.3 | 97.5±0.9 | 98.8±0.4 | 100±0.0 |
| 3M Aura 9322+ | New | 2 | 96.7±0.2 | 99.1±0.0 | 99.7±0.0 | 99.3±0.3 |
| | 1x | 2 | 91.0±1.0 | 99.0±0.0 | 100±0.0 | 100±0.0 |
| | 2x | 2 | 77.5±1.5 | 85.5±0.5 | 89.5±0.5 | 98.0±1.0 |
| San Huei 2920V | New | 2 | 95.5±0.5 | 99.0±0.0 | 100±0.0 | 100±0.0 |
| | 1x | 8 | 92.3±1.6 | 97.8±0.7 | 99.1±0.3 | 96.0±5.4 |
|  | 2x | 6 | 90.2±0.7 | 96.8±0.35 | 99.0±0.0 | 97.8±2.3 |
| Maco<br>Pharma<br>ZZM002 | 1x | 8 | 89.3±3.6 | 96.8±1.2 | 98.9±0.3 | 99.8±0.4 |
|  | 2x | 6 | 90.2±0.7 | 96.8±0.35 | 99.0±0.0 | 97.8±2.3 |
| Safe<br>Worker<br>1016 | New | 2 | 96.5±0.5 | 98.0±0.0 | 60.5±1.5 | 99±0.0 |
|  | 1x | 8 | 60.3±5.3 | 81.6±4.9 | 90.1±5.3 | 91.5±18.4 |

### Flow resistance

For the breathing resistance test we tested 6 FFP-2 respirators (3M Aura 1862+): two were used once and reprocessed, two were used twice and reprocessed after each use and two were used for 3 times and reprocessed after each use. The average pressure did not increase with the number of reuses (35.6±0.3%, 35.4±0.0%, 36.7±0.3%, respectively)

## Discussion

The COVID-19 pandemic has caused major shortages of PPE, including protective respirators. While production has increased, shortages are so high that reprocessing of used respirators and respirators is probably one of the only viable short-term solutions. In primary care, a tabletop autoclave would be a pragmatic choice, as the device is readily available in practices for sterilization of surgical and gynecological instruments. In this study we found that steam sterilization at 121 degrees Celsius may provide a viable option for selected respirators, but it also sheds a light on the variability in the protective properties of the various available respirators and respirators. Of the tested respirators, the filter capacity of the 3M Aura 1862+ respirator fared best with a consistently high filter capacity for the 0.3 μm particle size category and above after multiple cycles of steam sterilization. Moreover, there are no indications that the respirator becomes harder to breathe through and thus more uncomfortable to wear.

### Findings in relationship to FFP-2 and FFP-3 standards

The particle chamber used in this study appears to be more stringent (more sensitive) that the NEN-149 criteria that are used for FFP-2 and FFP-3 norms. We performed a cross-check with 4 KN95 respirators which showed that measurements of 67% and 82% particle retention at 0.3 and 0.5 microns on average, using our setup still resulted in approval for use according to the FFP-2 norm when measured according to the NEN-149, based on a continuous flow setup (Kalibra, Delft, the Netherlands). Therefore, apart from the Safe Worker 1016, all other mask types will likely still comply with NEN-149 FFP-2 threshold values.

### Study limitations

Our study involved a pragmatic study with a limited sample size of respirators and respirators available in our practice. Our study did not involve testing of surgical masks or FFP-1 masks, and we do not know whether reprocessing of these materials would still provide adequate protection based on their respective standards. Furthermore, we did not perform a laboratory-based Fit test. Also, we presumed that solid particles of 0.3-0.5 microns are of relevance and behave similar as droplets that normally carry viruses from one person to another. Moreover, we do not know exactly at what particle size viral transmission is still possible. Finally, this study did not test the efficacy of reprocessed respirators in a clinical trial setting, as such we do not know the “real world” safety of reprocessed respirators.

### Prior studies

In our primary care medical center, we have empirically experimented with decontamination of FFP-2 and FFP-3 respirators and respirators using steam sterilization using the medical autoclave. We found that when using steam sterilization at higher temperatures (132-140 centigrade), visual deformities occurred on the respirators, particularly those with a plastic respirator valve. A recent analysis of the Dutch Centers of Disease Control (RIVM) also found that steam sterilization at higher temperatures deformed the masks [12]. To our knowledge there is only one prior study that assessed the impact of steam sterilization on the filter capacity of facepiece respirators, which was a study by Lin and colleagues in 2017. [13] In this study, the authors found that one of the decontamination processes that appeared effective for N95 respirators was the medical autoclave, in which they exposed the respirators to saturated steam at 121 degrees Celsius for 15 minutes. They found that filter quality (≥95%) of the masks remained intact using a range of particles. These findings are comparable to those we present in this paper.

### Implications for practice

For COVID-19 like for other viruses, transmission can occur via droplets or aerosols. [1–4] Thus personal protection is warranted to avoid catching COVID-19. Currently there is no evidence on which type of face mask offers best protection for COVID-19. Prior studies with influenza viral particles showed that FFP2 respirators may provide better protection that surgical masks, when used appropriately. [14] In this COVID-19 pandemic, it is thought that the use of surgical masks may be sufficient for consultations with only limited person-to-person exposure. However, it is much less certain whether surgical masks will provide adequate protection during longer consultations or back-to-back consultations with patients with suspected COVID-19 in a closed consultation room. [15] In these instances, respirators may be preferable. Given, the limited availability, reusing FFP-2 type respirators may provide a second-best alternative that can be readily performed in primary care and other low-resource settings using a table-top medical autoclave, as described in this study.

## Conclusion

This study shows that selected FFP2 respirators may be reprocessed for use in primary care, as the respirators retain their shape, ability to retain particles and breathing comfort after decontamination using a medical autoclave. However, future studies are warranted to confirm our findings.

## Data Availability

All data will be made available on https://repository.tudelft.nl/

## Contributors

REH, BvS, BvM, JB, TH contributed to the study design and acquisition of data. REH, BvS and TH analyzed and interpreted the data. JB, JJvdD, JRMvdS contirbuted to the interpretation. REH drafted the initial manuscript and all authors critically revised the manuscript and gave final approval.

## Acknowledgements

The authors would like to thank Jan-Willem Klok and Tomas Lenssen for testing of all samples on the test setup.

## Funding statement

This work of research received no specific grant from any funding agency in the public, commercial or not-for-profit sectors.

## Competing interests

None declared.

## Patient consent for publication

Not required.

## Ethics approval

Not required.

## Data availability statement

All data will be made available on https://repository.tudelft.nl/

## Notes

### Competing Interest Statement

The authors have declared no competing interest.

